# Can dual active ingredient Interceptor® G2 insecticide treated net (ITN) replace indoor residual spraying (IRS) efficiently? A case study in Sakassou, Côte d’Ivoire

**DOI:** 10.64898/2025.12.22.25342668

**Authors:** Joseph Chabi, Gloria Salome Shirima, Brian Masanja, Sylvester Coleman, Constant Guy N’Guessan Gbalegba, Bernard Loukou Kouassi, Brice Renaud N’Guessan Broudje, Constant Victorien Ako Edi, Firmain N’Dri Yokoly, William Olatondji Adimi, Ruth-Marie Adjoua Kouame, Valentin Anian, Rosina Kyerematen, Alexander Egyir-Yawson, Samson Kiware, Samuel Kweku Dadzie

## Abstract

**Background:** Following three rounds of successful Indoor Residual Spraying (IRS) implementation in the district of Sakassou (Côte d’Ivoire), IRS was withdrawn and replaced by insecticide treated nets (ITNs). This study evaluated the entomological and epidemiological impacts of Interceptor (IG2) ITNs distributed in Sakassou, to determine if the protection offered by IG2 nets was adequate to suppress malaria transmission post IRS withdrawal.

**Methods:** The Vector Control Optimization Model (VCOM) was adapted to evaluate the effectiveness of the IRS and IG2 deployment on malaria transmission dynamics. Additionally, we used an interrupted time series (ITS) model to analyze routinely reported malaria cases in the district health Information Management System (DHIS2) to determine the epidemiological impact of both interventions. Counterfactual trends were generated for the post IRS withdrawal with IG2.

**Results:** The results of the VCOM showed a 55.4% (95%CI, 48.3 – 62.4) reduction in the human biting rate (HBR) when IRS was deployed and 48.8% [95%CI, 42.8 – 54.6]) when IG2 nets were distributed, compared to standard pyrethroid-only nets. No statistical difference was recorded between the HBR of IG2 and IRS (p=0.164). Similarly, there was no statistical difference between the entomological inoculation rate (EIR) of IRS (64.7% [95%CI, 56.6 – 72.8]) reduction and IG2 (61.9% [95%CI, 54.2 – 69.6]) reduction (p=0.616). Furthermore, the ITS showed 26% reduction in malaria cases that was recorded immediately after spraying (IRR = 0.74; 95% CI: 0.62–0.90; *p* = 0.002) with cumulative impact over time and spray rounds and similarly to IG2 performance (IRR = 1.00; *p* = 0.200).

**Conclusion:** The study findings suggest that IG2 nets provided comparable entomological efficacy as clothianidin-based IRS but could not adequately suppress malaria cases after IRS withdrawal which could be due to plastic vector feeding behaviour. This study provides evidence that dual AI ITNs such as IG2 may be warranted as a contingency strategy after IRS withdrawal but may require additional studies for full recommendation.

## Background

Malaria vector control has historically contributed the most to the declines in malaria burden in sub-Saharan Africa especially in the last two decades (1). In most endemic countries mass distribution of insecticide treated nets (ITNs) and indoor residual spraying of insecticides (IRS) (2, 3) have been the main vector control interventions used by the malaria programs. Since these interventions rely heavily on the efficacy of the insecticides used, vector resistance to the available public health insecticides have emerged as significant threat to their efficacy and has contributed to increasing malaria burden (4–7). It was reported that about 90 percent of the vectors in endemic countries were resistant to at least one of the pyrethroids used in the impregnation of ITNs (8, 9). In response to increasing insecticide resistance, malaria control programs have shifted to new-generation vector control products to maintain program efficacy. These include ITNs impregnated with an insecticide and a synergist, such as pyrethroid-piperonyl butoxide (PY-PBO), or with dual active ingredients, such as pyrethroid-chlorfenapyr (PY-CFP). Similarly, for IRS recently prequalified IRS products such as clothianidin-based insecticides (SumiShield and Fludora Fusion) and broflanilide (Vectron T500) have been introduced. However, this transition to these new products have huge financial implications because they are considerably more expensive than the previously used pyrethroid-only based products (10). It is estimated that IRS with the new insecticides cost about five times more each person protected per year compared with PY-PBO or PY-CFP ITNs (11). As a result, ITNs have been prioritized in most countries across Africa and are distributed widely than IRS. In places where IRS remains part of the malaria vector control strategy it is only selectively deployed in regions with the highest burden. This has affected the sustainability of IRS in many countries leading to reductions in the geographic coverage of IRS since 2010, facilitated by the decline in funding (12). Between 2017 and 2025, the number of countries supported by the U.S. President’s Malaria Initiative (one major IRS implementing partner), decreased from 17 to 6 countries (13). Despite the challenges with IRS, the strategy remains one of the best malaria vector control tools (14, 15). In places where IRS have been withdrawn or scaled back, its withdrawal has often been linked to a resurgence of malaria, with studies showing increased entomological indices of transmission as well as malaria incidence rates to pre-intervention levels (16–22).

The new generation of ITNs have been suggested as cost effective alternatives to IRS that can sustain the gains of IRS when it is withdrawn, but there is limited data to support this assumption. PBO-ITNs were the first new generation of ITNs incorporated with a synergist that enhances insecticide susceptibility of vector populations introduced. A meta-analysis on their efficacy in Africa, predicted that compared to pyrethroid-only (PY) ITNs, the PBO ITNs could avert up 501 (95%CI 319–621) cases per 1000 people per year in places where there was high pyrethroid resistance(4). However, some studies have shown that despite their efficacy, the effectiveness of PBO ITNs often waned significantly after two years, even though they are designed to last three years (23, 24) Interceptor® G2 (IG2) a first-in-class dual active ingredient ITN that combines two different insecticides (chlorfenapyr & alpha-cypermethrin) were also recently prequalified (25). Studies have shown remarkable efficacy of IG2 ITNs against malaria transmission, with a much-sustained efficacy against malaria incidence over the duration of its 3-year life span when compared to PBO ITNs (26–30). The overall performance of IG2 has led to it being recommended for use in many countries challenged by high pyrethroid resistance and as a replacement for IRS.

In Côte d’Ivoire, the NMCP and partners supported the implementation of three consecutive IRS campaigns from 2020 to 2022 recording a remarkable reduction of malaria transmission in an area with intense pyrethroid resistance (31, 32) (33–35). Nonetheless, the strategy had to be discontinued due to the cost of implementation and the need to prioritize resources to fund the scale up of new generation ITNs across the country to protect the entire population at risk. Following the IRS withdrawal, the population of Sakassou were provided with IG2 ITNs. This decision was primarily based on the entomological efficacy of IG2 demonstrated in semi-field trials (28, 36). Though there is limited data to suggest that IG2 can provide the protection needed when IRS is withdrawn from an area where IRS has had significant impact on malaria transmission and cases. This study was therefore conducted to assess the entomological and epidemiological impact of IG2 nets in an IRS withdrawn district and determine whether IG2 could suppress malaria transmission following the withdrawal of IRS in a highly endemic setting.

## Methods

### Study site

The study district was Sakassou, situated in the pre-forested savannah zone and has a surface area of approximately 1,820 km^2^. The climate is tropical with two rainy seasons (March to June and September to October) and two dry seasons (July to August and November to February). However, in recent years, there have been variations in rainfall due to climate change. The average annual rainfall is about 899.6 mm, and the mean annual temperature is 26°C. Sakassou district has 172 villages and hamlets, and 28 health facilities that serve a population of about 126,470 inhabitants (Fig. 1). The district recorded the highest malaria incidence in Côte d’Ivoire between 2015 and 2020 according to the NMCP (37).

**Fig. 1:**
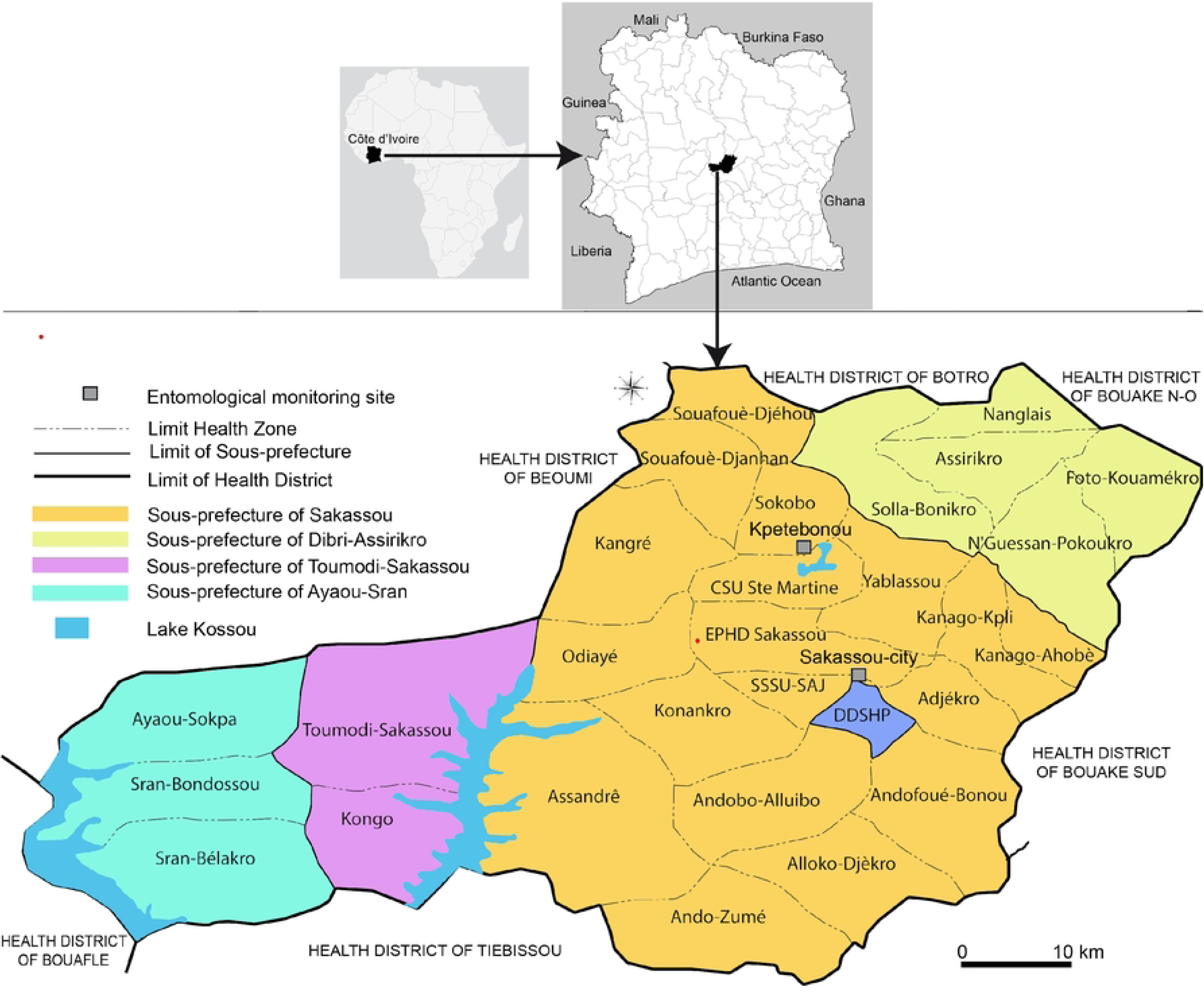
Map of the district of Sakassou showing the two entomological monitoring sites and 28 heath zones from where epidemiological data were collected.

### Vector Control Interventions

Within the NMCP’s national strategic plan (NSP) of the period 2016-2021, the country conducted its third ITN mass distribution in 2017 where PY-ITNs were distributed in Sakassou. Afterwards, IRS was implemented once a year from 2020 to 2022 with an overall mean coverage of 94.6% and 128,817 inhabitants protected (32) over the three consecutive years of implementation. The 2023 mini-ITN distribution campaign post IRS withdrawal specifically conducted for the IRS sites achieved high operational coverage in Sakassou, with 96% of planned ITNs distributed (38). Overall population coverage reached 93%, and 97% of households received at least one ITN, indicating near-universal access following the campaign.

### Entomological data collection

Human landing catches (HLCs) and pyrethrum spray catches (PSCs) were conducted every month in Sakassou before during and after IRS and introduction of IG2 from January 2019 to December 2024. HLCs were carried out in four houses and PSCs in 30 houses during two consecutive days each month to capture any seasonality related to vector density and behaviour. Mosquito collections were conducted in the same houses throughout the study period.

HLCs were performed inside and outside of the four houses from 6:00 p.m. through 6:00 a.m. A team of sixteen volunteer mosquito collectors from whom written consents were received (two teams of 8 mosquito collectors, each working half night shifts) conducted HLCs every month.

All mosquitoes collected through each method were morphologically identified to genus. *Anopheles* mosquitoes were identified to species or species complex by binocular microscope, using identification keys (39). All malaria vectors were preserved on silica gel in Eppendorf tubes and processed in the laboratory to identify sibling species and determine *Plasmodium* infection status. Genomic DNA was extracted from the legs and wings of individual anopheles mosquitoes using the LIVAK method (40) and processed by polymerase chain reactions (PCR). Members of the *An. gambiae* s.l. complex species (*An. gambiae* s.s., *An. coluzzii* and *An. arabiensis*) were identified by SINE-PCR (41).

To detect the presence of *Plasmodium* parasites, the head and thorax of *An. gambiae* s.l, and *An. funestus* s.l. collected using HLC were analyzed by enzyme-linked immunosorbent assay (ELISA) to detect circumsporozoite proteins as described by Wirtz et al, (42). This method uses a monoclonal anti-body that recognizes a repetitive epitope on the circumsporozoite protein of *Plasmodium falciparum*. ELISA Reagent Kits (MRA-890) were obtained from BEI Resources (NIAID, NIH, USA). Examination of assayed samples was done after reading optical densities (OD) at 405 nm on an ELISA plate reader (Biotek ELx800, Swindon, UK). Positive samples were determined by OD readings 2-fold greater than the negative controls and were tested a second time for validation of all positive samples detected during the first test using the boiling method (43).

### Epidemiological data collection

From each of the 28 health facilities (Figure 1), monthly confirmed malaria cases was retrieved from the health management information system (HMIS). Health facility data was extracted from consultation registers at all health facilities of the district. For each patient recorded in the registers, the RDT result and microscopy result were recorded for the period of the study. Confirmed malaria case was defined as any positive RDT or microscopy result.

## Statistical analysis and data interpretation

### Modelling Entomological impact of the different interventions

The Vector Control Optimization Model (VCOM) (44), a computational tool used to predict the impact of combined vector control interventions on mosquito populations was used to evaluate the effectiveness of the ITNs and IRS deployment on malaria transmission dynamics (44). Historical data from Sakassou was used to estimate the intervention specific parameters, while other mosquito transmission parameters of the *An. gambiae* s.l. were adapted from the VCOM model described by (44). Simulations were performed using field data to assess the field scenario and also generate predictions on the continued deployment of a given intervention in order to assess its impact over time.

The model simulated the effects of three primary interventions: first, standard PY-ITNs implemented prior to the introduction of IRS from January 2019 to July 2020. Secondly, IRS conducted using clothianidin-based insecticides from August 2020 through July 2023 followed by, IG2 implemented in August 2023 and data through December 2024. The intervention timelines in the model simulations were designed to mimic implementation scenarios with specific months designated for the initiation and cessation of each intervention. The main entomological outcome indices used to assess the entomological efficacy of these interventions were the indoor resting density (IRD = mean number of female mosquitoes per room), human biting rate (HBR = mean number bites per person per night) and the entomological inoculation rate (EIR = mean infected bites per person per night).

### Epidemiological data and predictions

An interrupted time series (ITS) analysis was used to assess the immediate and sustained post-IRS effect as well as the effect of IRS withdrawal on confirmed malaria cases in Sakassou. A negative binomial regression model with time as a continuous variable, month as a categorical variable to adjust for seasonality, and a binary indicator for the IRS period was fitted. Malaria case ratios (IRRs) and 95% confidence intervals (CIs) were calculated for the intervention variable (IRS) to estimate the immediate change following IRS introduction. Monthly dummy variables controlled for seasonal malaria transmission patterns. A separate model was fitted for a secondary exposure variable (IG2), representing a different intervention or analysis scenario. All analyses were performed using R 4.5.2 software.

## Results

### Trends in entomological parameters

#### Indoor resting density (IRD)

The analysis from the VCOM showed that IRS resulted in an immediate 52.0% (95% CI 46.5 - 61.8) reduction in the IRD of the predominant vector when compared to the PY-ITN period, which was sustained through the period of IRS implementation (Table 1). In comparison to the PY-ITN (pre-IRS) period, IG2 also resulted in about 45.0% (39.9 - 54.2, 95%CI) reduction in IRDs (Fig. 2, Supp. data 1). The impact of IRS and IG2 ITN deployment on the IRD of *An. gambiae* s.l. was comparable (p = 0169) considering the same period (Table 1).

**Fig. 2:**
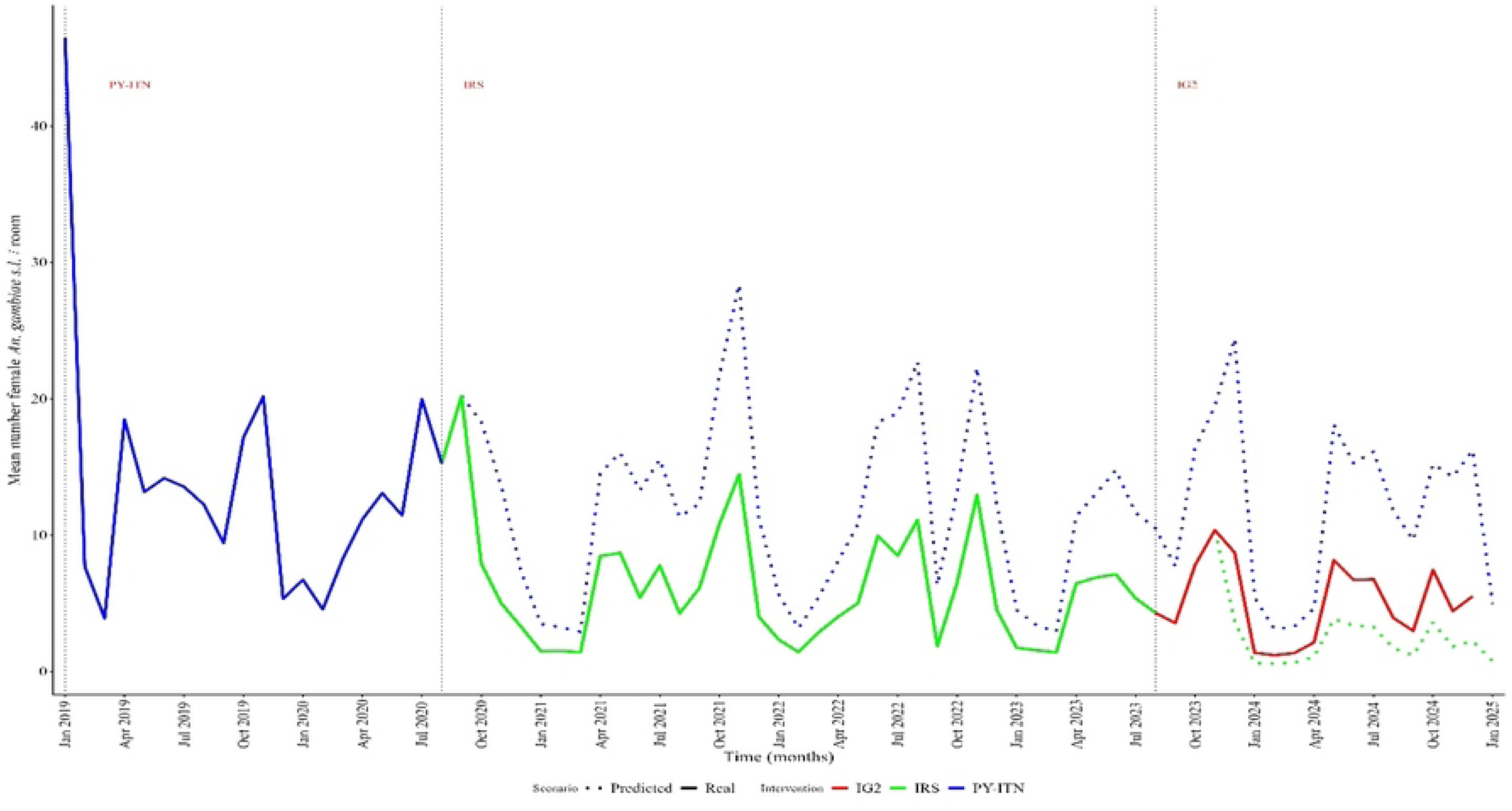
Observed and predicted impact of IRS and IG2 ITNs on the mean IRD of An. Gambiae s.l. from January. 2019 **to December** 2024. The vertical dotted lines delineate the different intervention periods. PY-ITN Implementation: Shown in blue (solid line), indicating the IRD during PY-ITN coverage. The blue dotted line represents predictions that the IRD had for PY-ITN distribution continued over time. IRS Implementation: Represented in green (solid line) with a dashed green line, highlighting the predicted impact of IRS on mosquito populations during the spray intervention. *IG2: Shown in red, reflecting the outcomes of this separate intervention and its impact over the years*.

**Table 1:**
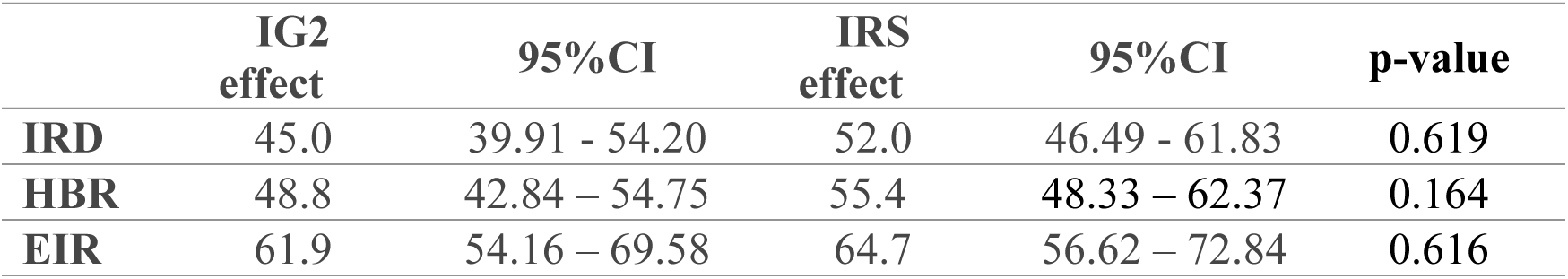
Overall impact of IRS and IG2 on entomological indices highlighting the similarity of the entomological indices during both interventions

### Human biting rate (HBR)

Simulation from the model indicates a greater reduction in human biting rates during the IRS period (55.4% [95%CI, 48.3 – 62.4]) compared to when IG2 nets were deployed (48.8% [95%CI, 42.8 – 54.7]) with the standard PY-ITNs (Table 1). A counterfactual scenario of continued IRS would have resulted in a further decline in HBR than was observed when IG2 were deployed. However, this projected IRS impact was comparable to the observed effect that the IG2 ITNs had on the HBR of *An. gambiae* s.l. (p=0.164) (Fig. 3, Table 1, Supp. data 1).

**Fig. 3:**
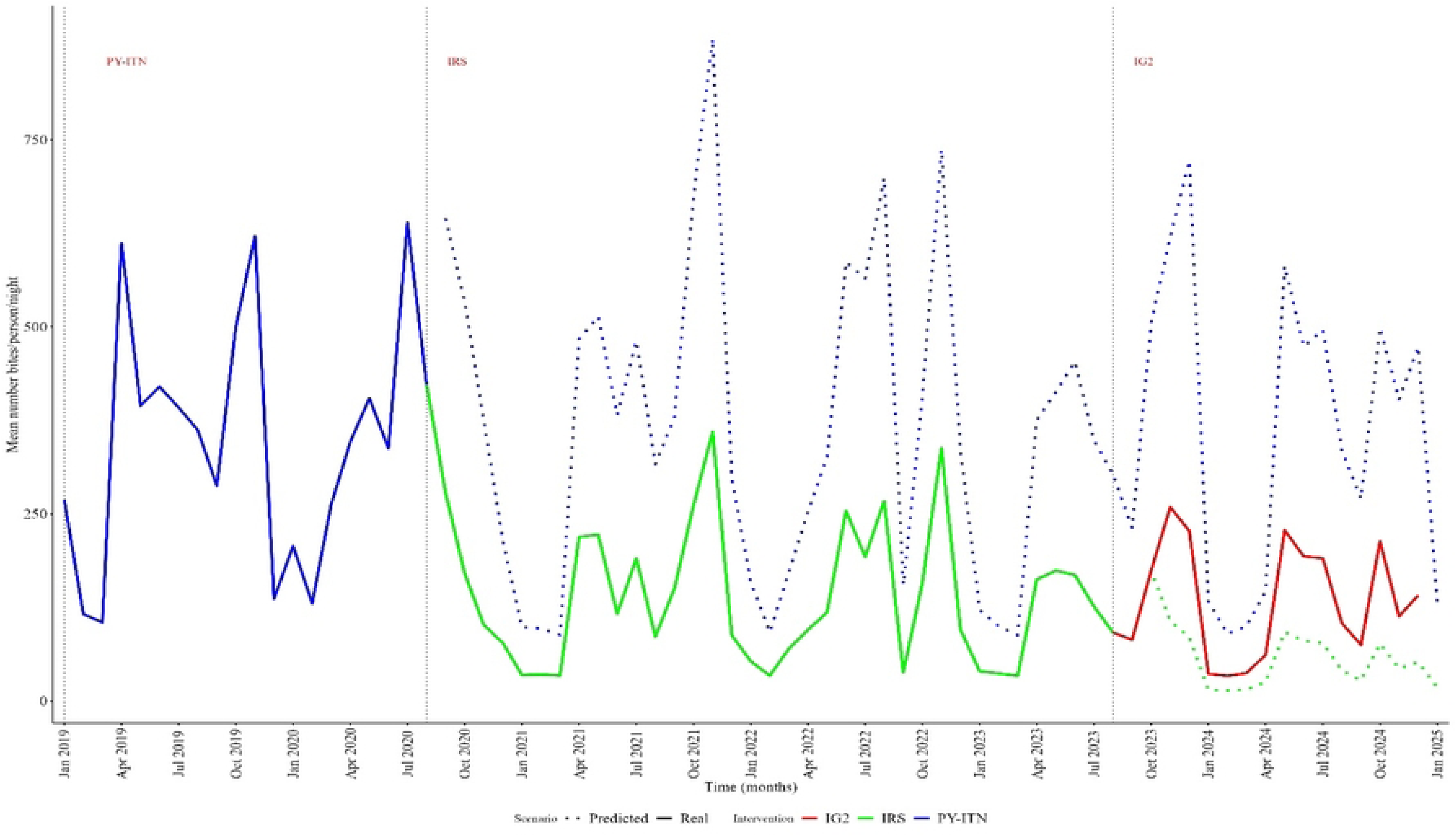
Observed and predicted impact of IRS and IG2 ITNs on mean HBR of An. Gambiae s.l. from January. 2019 **to December** 2024. *The vertical dotted lines delineate the different intervention periods. PY-ITN Implementation: Shown in blue (solid line), indicating the HBR An. gambiae s.l. mosquitoes during PY-ITN coverage. The blue dotted line represents predictions that the HBR had for PY-ITN distribution continued over time. IRS Implementation: Represented in green (solid line) with a dashed green line, highlighting the predicted impact of IRS on the HBR during the spray intervention*. *IG2: Shown in red, reflecting the outcomes of this separate intervention and its impact over the years*.

### Entomological inoculation rate (EIR)

Figure 4 shows observed and predicted EIRs when PY-ITN, IRS or IG2 was deployed. The analysis indicates that both IRS and IG2 resulted in significant declines in EIR when compared to the baseline period when PY-ITNs were used. IRS resulted in a 64.7% (95%CI, 56.6 – 72.8) decline in EIR whilst IG2 resulted in a comparable reduction of 61.9% (95%CI, 54.2 – 69.6 [p=0.616]) over the same period (Fig. 4, Table 1, Supp. data 1).

**Fig. 4:**
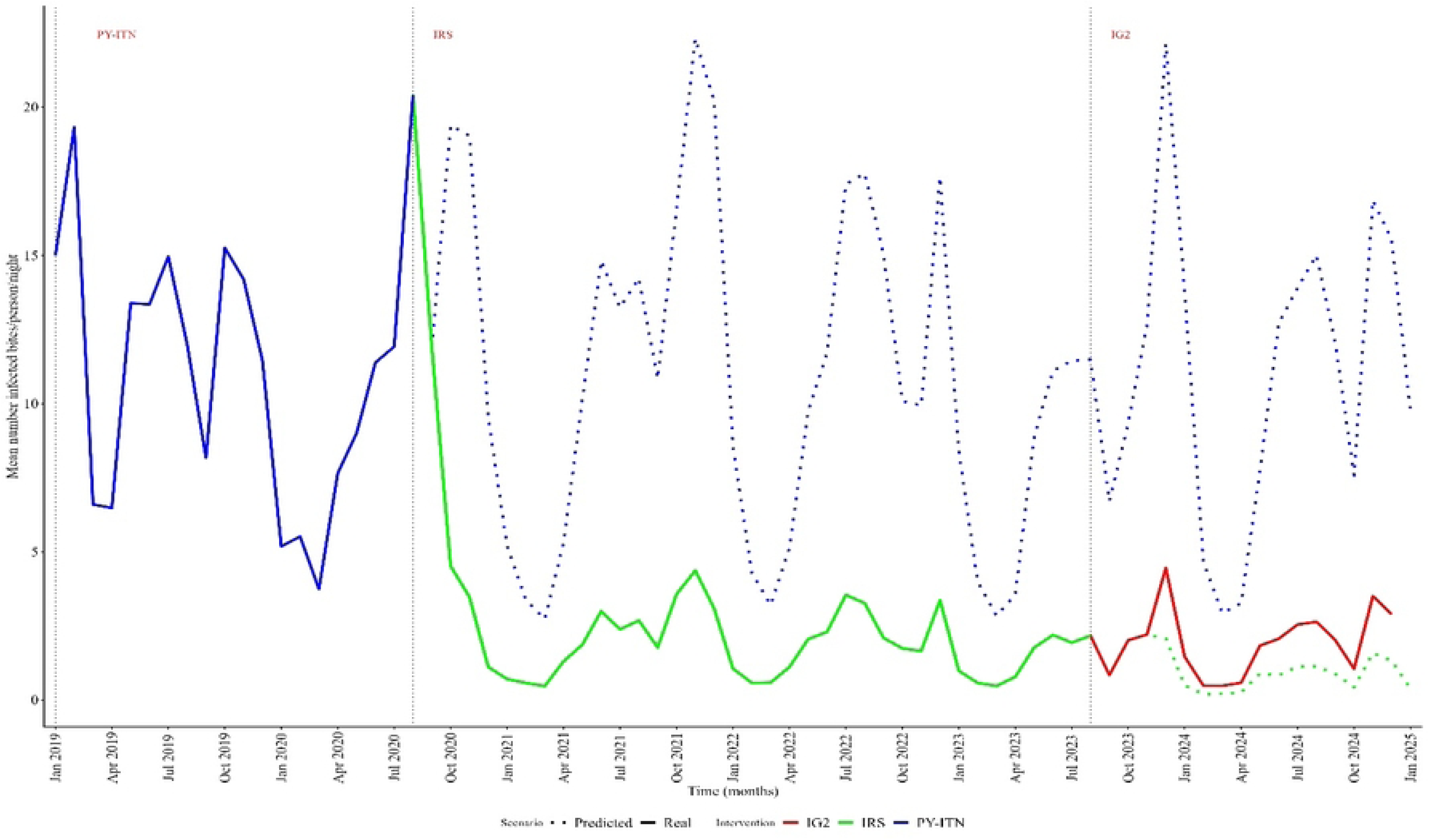
Observed and predicted impact of IRS and IG2 ITNs on mean EIR of *An. gambiae* s.l. from January. 2019 **to December** 2024. The vertical dotted lines delineate the different intervention periods. *PY-ITN Implementation: Shown in blue (solid line), indicating the EIR An. gambiae s.l. mosquitoes during PY-ITN coverage. The blue dotted line represents predictions that the EIR had for PY-ITN distribution continued over time. IRS Implementation: Represented in green (solid line) with a dashed green line, highlighting the predicted impact of IRS on the EIR during the spray intervention*. *IG2: Shown in red, reflecting the outcomes of this separate intervention and its impact over the years*.

## Trends in epidemiological parameters

### Effect of IRS on the number of malaria cases

The ITS observational analysis showed that IRS implementation in Sakassou was associated with a 26% reduction in malaria cases immediately after spraying (IRR = 0.74; 95% CI: 0.62–0.90; *p* < 0.001). The underlying time trend showed a small but statistically significant increase in malaria cases over time (IRR = 1.01; 95% CI: 1.00–1.01; *p* < 0.001).

Monthly analysis demonstrated marked reductions in malaria cases in February (IRR = 0.68; *p* = 0.003), March (IRR = 0.65; *p* < 0.001), and December (IRR = 0.73; *p* = 0.012) compared to January, highlighting a seasonal pattern of transmission. The number of malaria cases in other months remained largely unchanged (IRRs ≈ 0.9–1.0, p > 0.05).

### Effect of IRS withdrawal

A modelled counterfactual scenario of the withdrawal of IRS and re-introduction of PY- ITNs in Sakassou was associated with a statistically significant surge in malaria cases. The Rate Ratio of the number of confirmed malaria cases during post-IRS withdrawal period was 1.34 (95% CI: 1.08–1.67, *p* = 0.009), indicating a 34% increase compared to the counterfactual scenario of sustained IRS intervention (Fig. 5).

**Fig. 5:**
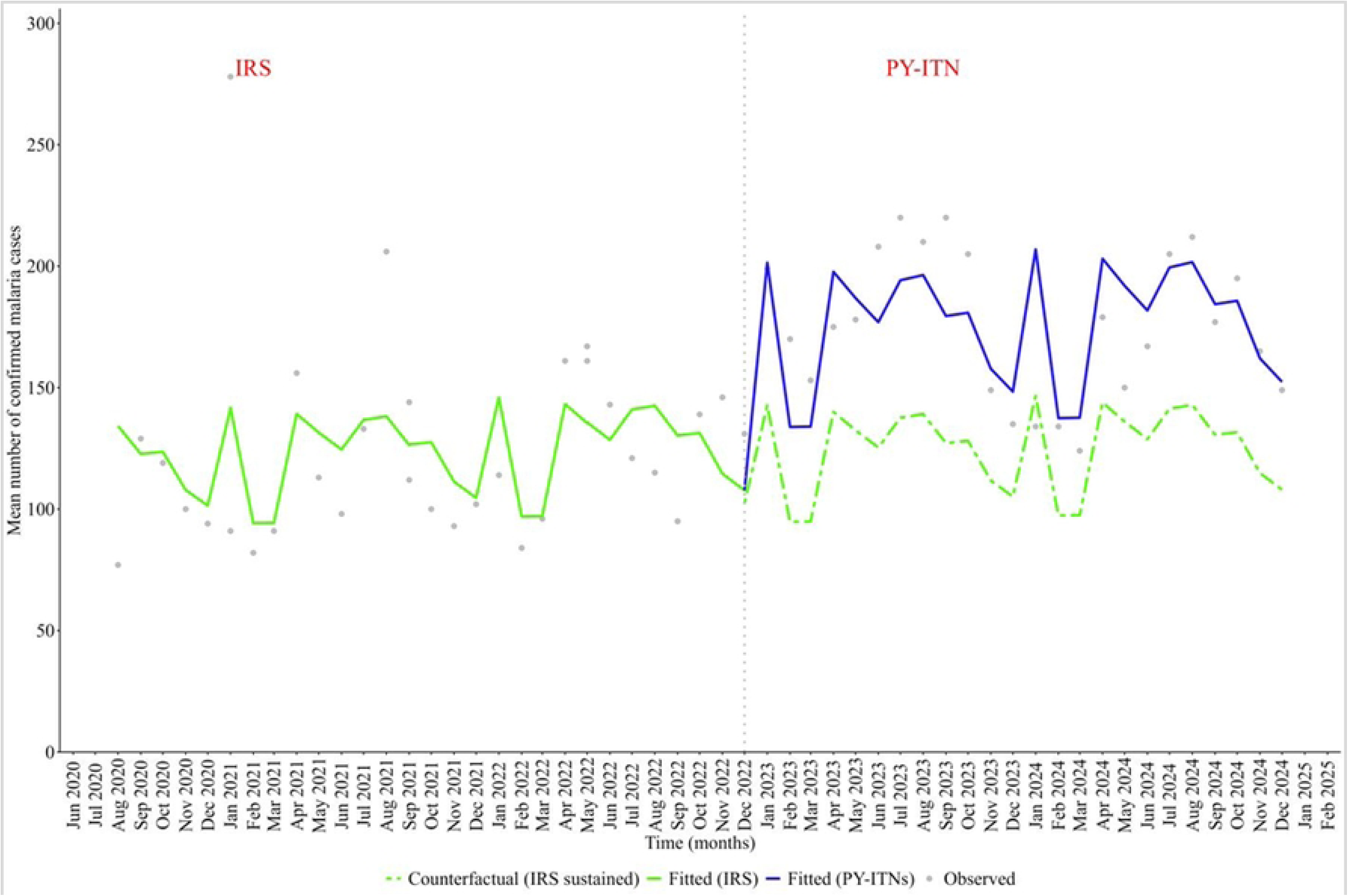
**Observed and counterfactual number of malaria cases in Sakassou during the period of IRS implementation and IRS withdrawal**. The vertical dotted line graph delineates the potential end of effect post-IRS implementation. *IRS and PY-ITN implementation: Shown in green (solid line), indicating the number of malaria cases during IRS coverage. The green dotted line represents predictions that the number of malaria cases might have been during IRS Implementation. IRS withdrawal effect represented in blue (solid line) highlighting the predicted impact of IRS withdrawal on the number of malaria cases over the subsequent year*.

### Secondary model (IG2 treatment variable)

When data on IG2 was included in the model, there was a marginal increase of about 16 % in malaria cases following IRS withdrawal, however this was not statistically significant (IRR = 1.16; 95% CI: 0.97–1.40; *p* = 0.120) when compared to IRS during the same period (Table2). Seasonal reductions were evident during the dry months of February (IRR = 0.71; *p* = 0.011), March (IRR = 0.68; *p* = 0.004), and December (IRR = 0.75; *p* = 0.027). The time trend on malaria cases was not statistically significant (IRR = 1.00; *p* = 0.20), suggesting a sustained impact of IG2 over the period (Fig. 6, Supp. data 2).

**Fig. 6:**
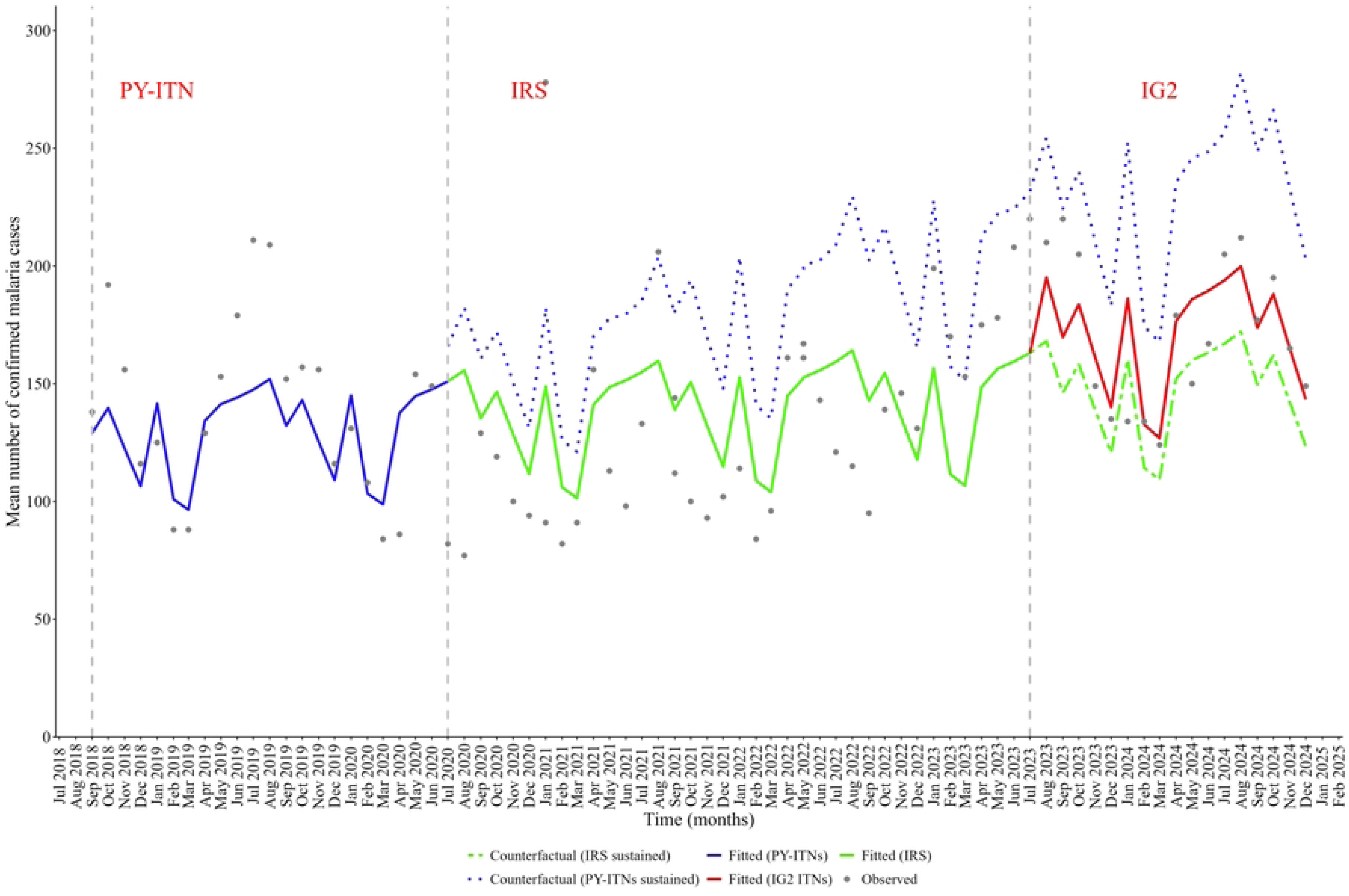
**Observed and counterfactual number of malaria cases in Sakassou prior to the introduction of IRS, during the period of IRS implementation and post-IG2 introduction (after IRS withdrawal)**. *The vertical dotted lines delineate the different intervention periods. PY-ITN Implementation: Shown in blue (solid line), indicating the number of malaria cases during PY-ITN coverage. The blue dotted line represents predictions that the number of malaria cases might have been for PY-ITN distribution continued over time. IRS Implementation: Represented in deep green (solid line) with a dashed green line, highlighting the predicted impact of IRS on the number of malaria cases during the spray intervention*. *IG2: Shown in red, reflecting the outcomes of this separate intervention and its impact over the years*.

## Discussion

This study evaluated the performance of IG2 ITNs in sustaining malaria vector control gains in Sakassou district, Côte d’Ivoire, after IRS was discontinued. Historically Sakassou has been known to be a high burden malaria endemic district that contributes the most to the malaria burden in Côte d’Ivoire, with transmission sustained mainly by *An. coluzzii*. Implementation of the IRS with clothianidin-based insecticides resulted in a steady decline in malaria incidence from 15.9% one year after IRS to about 38% by the end of the third round of IRS (31). This study was designed to determine if IG2 nets could sustain the reductions in malaria transmission indices and suppress malaria cases witnessed during the three rounds of IRS (31, 32). The findings suggest that IG2 nets provided comparable entomological efficacy as IRS but could not adequately suppress malaria cases after IRS withdrawal.

Even though IRS was effective to drastically decrease malaria transmission and incidences the challenge of cost in the implementation of IRS across malaria endemic countries is continuously affecting the number of countries that are able to sustain the strategy (15, 45–49). The study showed that continuing the implementation of IRS cumulatively reduced malaria vector density and malaria cases in the study area. The observed prediction of five-year IRS implementation reduced both entomological and epidemiological indicators of malaria in Sakassou. The data analysis showed that the additional years of implementing IRS suppressed the vector density up to 70% and this resulted in an immediate and significant reduction of approximately 26% of malaria cases in Sakassou. This is consistent with previous studies that showed that IRS was an effective vector control intervention and the trend is confirmed by several reports from countries like Ghana, Benin and Uganda where IRS was conducted for over a decade (15, 47, 50–52).

However, the marginal increase (16%) in the number of malaria cases 15 months post- IG2 distribution, though non-significant, contrasts with findings from other studies in West and East Africa. In Ghana and in Tanzania deployment of IG2 ITNs was associated with about 30% and 44% reduction in malaria incidence respectively (23, 53). Considering that IG2 reduced vector biting and transmission intensity, the apparent lack of epidemiological impact observed here may therefore reflect factors unrelated to the intrinsic bioefficacy of IG2 ITNs. The hypothesis is that this could be due to protection gaps resulting from possible human and vector behaviour patterns that allow significant human–vector interaction beyond the reach of IG2 nets (54–56), or operational decisions that resulted in sub-optimal protection.

*An. coluzzii* was reported feeding equally indoors and outdoors in Sakassou (32). This plastic feeding or a gradual shift to outdoor biting after three rounds of IRS (with deltamethrin and clothianidin), coupled with human night-time outdoor activities, may have led to significant exposure even when people were presumed protected by ITNs (57, 58). Increased insecticide pressure from the scale up of ITNs has been shown to increase exophagy and exophily (59) and this can sustain malaria transmission outdoors (60–62). Findings from the previous semi-field trials in M’bé, Côte d’Ivoire showed that IG2 ITNs induced deterrence and exit rates up to about 50%. In Tanzania and Uganda, 12% and 49% of the malaria transmission respectively occurred before sleeping time (63). Additionally, in places where individuals spend significant times outdoors between evening and early morning, either sleeping outdoors (without a ITNs) or involved in night-time outdoor activities without any personal protection, there is a high risk of acquiring malaria (64–68). In such instances additional messaging on correct and sustained use as well as the use of additional personal protection and larval source management may be warranted to complement indoor targeted interventions. Additionally, the timing of the deployment of vector control interventions also plays a key role in the epidemiological impact of the intervention. The optimal time in the year for deployment of interventions is before the transmission season (69).

Our analysis indicate that malaria transmission is perennial in Sakassou and highly variable by month, with peaks that typically coincides with the long rainy season (March-July) and the end of the short rainy season (September-November) (70). Therefore, the optimal timing for deployment of ITNs would have been in March/April 2023. However, the IG2 TNs were distributed in August 2023 during the peak of the malaria transmission season starting from July 2023. Also, there was about 15 months between the last IRS campaign in May 2022 and the distribution of IG2 in August 2023, which could have also affected the trends in malaria cases. By March 2023, 10 months after the third year’s IRS which is beyond the residual life of the sprayed insecticide (SumiShield), we had begun to see a slight rise in malaria cases from (71–73). Starting interventions late into the transmission season do not only increase the risk of widespread transmission but also amplifies the infectious reservoir, resulting in sub optimal protection. Findings from a study in Wajir County in Northeast Kenya highlights this risk and confirms that the timing of vector-control interventions is critical in determining malaria outcomes in settings where malaria transmission is seasonal (74). The authors reported that delayed deployment of vector control interventions (six months after heavy rainfall and flooding) in Wajir county in 1998 was associated with a large and explosive malaria epidemic (with weekly incidence rates peaking at 54/1,000 population/week). In contrast, no epidemic was recorded in 2007 when vector control interventions were deployed within three months of the start of the rainy season. Weekly malaria incidence rates never exceeded 0.5 per 1,000 population per week. The deployment of IG2 ITNs at the peak of the malaria transmission season in our study district might account for the marginal increase in the number of malaria cases.

Despite that marginal increase in malaria cases following IRS withdrawal, IG2 deployment appears far more cost-effective than the counterfactual scenario of deploying PY-only ITNs, which would led to a 34% increase in cases as previously reported in several countries where IRS was stopped (16, 75, 76). Conversely, additional years of implementing IRS deployed at the right time could have suppressed the vector density up to 70% and which could have resulted in additional 26% reduction in malaria cases in Sakassou. This is consistent with previous studies from Benin, Ghana and Uganda (where IRS was conducted for over a decade) (15, 47, 50–52). These studies showed that repeated IRS with the right insecticide deployed at the right time could result in significant and sustained entomological and epidemiological impacts.

However, operational adjustments such as improving timing and user education may result in improving the epidemiological impact that was expected. Still, it will be important to continue monitoring the impact of IG2 over time, and additional studies in different malaria transmission settings will be required to generate further evidence on whether IG2 can replace IRS or help sustain the gains.

## Limitations

The IG2 model may require additional parameters that were not included in the model and suggests that either the intervention variable did not capture a true change in exposure or that other confounding factors such as population behaviour related to ITN use or vector’s attitude towards the insecticide in the ITNs, malaria case management and commodities stock outs may have diluted the signal. Further investigations are needed to clarify whether this represents a true null effect or challenge with data/measurement.

## Conclusion

The study reports that replacing IRS with IG2 nets was associated with a marginal but not significant increase in the number of malaria cases in Sakassou and similarly to the malaria transmission indicators such as the IRD, HBR and EIR. Overall, the study shows how the dynamics of malaria transmission and operational decisions could impact the effectiveness of both IRS and ITNs as vector control tools. These findings presents key information for malaria programs and policy makers for consideration when deploying vector control interventions as countries work towards malaria elimination.

## Data Availability

All data produced in the present study are available upon reasonable request to the authors

## Acknowledgements

We are grateful to all site mosquito collectors and administrative authorities for facilitating the field data collections.

## Availability of data and materials

The datasets used and/or analyzed during the current study are available in the supplementary data set and could also be provided by the corresponding author on reasonable request.

## Abbreviations

An.: Anopheles
CI: Confidence interval
DHIS2: District Health Information Software version 2
EIR: Entomological Inoculation Rate
ELISA: Enzyme-Linked Immunosorbent Assay
HBR: Human Biting rate
HLC: Human Landing Catch
ITN: Insecticide Treated Net
IRD: Indoor Resting Density
IRS: Indoor Residual Spraying
NMCP: National Malaria Control Programme
PBO: Piperonyl Butoxide
PCR: Polymerase Chain Reaction
PSC: Pyrethrum Spray Catch
PY: Pyrethroid
RR: Rate Ratio
WHO: World Health Organization

## Authors’ contributions

JC^#^ (the corresponding author) organized the data set and drafted the manuscript. GSS, BM, SC and SK conducted data analysis and modeling. GCNG, BLK, BRNB, CVAE, FNY, WOA, RMAK and AV supported field collections, and reviewed the manuscript. BRNB supported data digitalization, data entry, cleaning and export. SC, GCNG, RK, AEY, AKD and SKD revised the manuscript for improvement. All corresponding authors reviewed the draft, provided inputs, which were collated and incorporated by JC. All authors read and approved the final manuscript.

## Declarations

The findings and conclusions expressed herein are those of the authors

## Ethical considerations

The surveys were conducted in accordance with the study protocol approved by the National Ethics Committee for Life and Health Sciences (CNESVS) of the Ministry of Health and Public Hygiene under the registered number 045-20/MSHP/CNESVS-kp. Written informed consent was obtained from household leader before selecting the houses as collection site. Volunteers were required to sign a consent form before participating in the study.

**Consent for publication** Not applicable **Competing interests**

The authors declare that they have no competing interests.

## Additional files

**Supplementary data 1:** All intervention malaria transmission indicators

**Supplementary data 2:** Malaria cases of all health facilities of the district

